# Prevalence of IgG and IgM antibodies to SARS-CoV-2 among clinic staff and patients

**DOI:** 10.1101/2020.07.02.20145441

**Authors:** Marcus Inyama Asuquo, Emmanuel Effa, Akaninyene Otu, Okokon Ita, Ubong Udoh, Victor Umoh, Oluwabukola Gbotosho, Anthonia Ikpeme, Soter Ameh, William Egbe, Margaret Etok, Jochen Guck, Andrew Ekpenyong

## Abstract

The coronavirus disease 2019 (COVID-19) is now a pandemic with devastating social and economic consequences. The extent of the spread of COVID-19 within populations is uncertain since diagnostic tests have not been carried out on all eligible persons and doing such diagnostic tests on everyone is much less feasible in developing countries such as Nigeria. Tests for antibodies to SARS-CoV-2, the virus that causes COVID-19, are more affordable, readily available, and require minimal training than current diagnostic tests. Employing a seroepidemiological strategy, serological tests were conducted on 66 volunteering staff and patients at the University of Calabar Teaching Hospital (UCTH), a Federal Government owned tertiary healthcare facility, to determine the extent of exposure to SARS-CoV-2, from 17^th^ to 25^th^ June 2020. Using a COVID-19 IgG/IgM Rapid Test Cassette with emergency use authorization (EUA) from the Food and Drug Administration (FDA) of the United States, it was observed that of the 66 samples tested, 5 (7.6%) were both IgG and IgM positive and 17 (26%) were IgG positive. Moreover, for 44 of the 66 participants, simultaneous tests were carried out using a rapid test kit from a different manufacturer but without FDA-EUA and all the results completely matched with the FDA-EUA kit, except one case where the FDA-EUA kit showed positive for both IgG and IgM while the other kit was positive only for IgM. The 26% positive IgG indicates a high exposure rate for the hospital staff and patients and points to community transmission where the facility is situated. Hence, immediate activation of WHO guidelines for controlling community transmission is called for. These results can further serve as a pilot study to guide public health policies in response to COVID-19 pandemic in both the general population and in healthcare settings.

## 1. INTRODUCTION AND BACKGROUND

The outbreak of the corona virus disease COVID-19 caused by a novel virus called SARS-CoV-2 Coronavirus^1–3^ has led to a pandemic^4^ that calls for both immediate and long term responses. Immediate responses have included large scale diagnostic testing using assays based on real-time reverse transcriptase polymerase chain reaction^5–8^ (RT-PCR) carried out in laboratory settings on respiratory specimens. As a nucleic acid amplification technique where RNA is converted into DNA and repeatedly multiplied for detection, RT-PCR, is currently the reference standard for diagnosis of COVID-19^5,9^. Cheng and colleagues provide a narrative review of challenges surrounding diagnostic testing for SARS-CoV-2, challenges largely occasioned by the nature of RT-PCR^5^. They note that many affluent countries have encountered problems with test kit contamination, accuracy, test delivery, and specimen collection that have inhibited rapid increases in testing capacity and that such challenges are even greater in low resource settings^5^. For instance, access to the primers and other supplies in the detection workflow remains a serious drawback for a developing country such as Nigeria. Furthermore, variations in primers adopted by different PCR test kit producers can add to the issue of access for developing countries that rely on vendors from advanced economies. For instance, the WHO-backed assay has primer-probe sets that target the SARS-CoV-2 RNA-dependent RNA polymerase (RdRP) and envelope (E) genes^5^ while the US uses its CDC-developed assay with PCR primer–probe sets for 2 regions of the viral nucleocapsid gene (N1 and N2) and for the human RNase P gene^5,10^.

Besides RT-PCR-based diagnostic tests which typically require special laboratory settings, there are moderately complicated molecular based assays that are faster than RT-PCR and are aimed at point-of-care usage such as the Abbott ID NOW COVID-19 (Abbott Laboratories)^11,12^ and the Xpert Xpress SARS–CoV-2 test (Cepheid)^13^ both of which have received the US Food and Drug Administration (FDA) emergency use authorization. The Abbott ID NOW COVID-19 which can produce results in 5 minutes can also produce up to 15% false negatives^14^ (if viral transport media are added to samples), highlighting a major limitation of current point-of-care molecular assays for SARS-CoV-2 clinical diagnosis. At the lowest rung of the ladder of complexity for testing are antibodies or serologic tests.

Serologic tests are carried out on clinical specimens such as blood, saliva, or even tears. COVID-19 serologic assays identify antibodies to SARS-CoV-2, such as IgA, IgM, and IgG. They are often based on enzyme-linked immunosorbent assays which are less complex than molecular tests^5^. Since they cannot rule out the presence of a virus, serology tests are of limited value in the immediate clinical diagnosis of a patient with suspected COVID-19 infection. Abbasi discusses both the promise and peril of antibody testing for COVID-19, calling for more oversight from regulatory bodies, such as the FDA and CDC, anchored on emerging scientific understanding of SARS-COV-2 and the pathophysiology of COVID-19^15^. However, positive results from validated antibody tests designed to be very specific to the SARS-CoV-2 virus can indicate whether a patient has COVID-19 infection, for IgM antibodies, or whether a patient has likely recovered from COVID-19 infection, for IgG antibodies^16^. The US Centers for Disease Control and Prevention (CDC) indicates an important usefulness of serological tests which captures the essence of our work, namely, that serologic tests are helpful in understanding how many people have been infected or exposed and how far the pandemic has progressed^16^. Although still being studied and debated^17^ owing to the novelty of COVID-19, an advantage of serological tests for COVID-19 is that the antibody tests could reflect immunity from COVID-19 and guide decisions on when to relax social distancing measures. Another possible advantage is that the presence of antibodies in recovering patients may signal the clinical utility of appropriate convalescent plasma for treating infected patients^18^. Beyond possibilities, compared to PCR and diagnostic molecular tests, antibody tests are more affordable and require minimal training for use.

With about 4-7% fatality^2,3^ and up to 20% hospitalization^2^ in some countries, the pandemic is a serious challenge to the human family and the economy. Immediate efforts to prevent or reduce transmission through non-pharmaceutical interventions such as hand washing, respiratory hygiene, wearing of face masks, social distancing, ban on large gatherings, border closures, movement restrictions, school closures, partial lockdown, and total lockdown, have already occasioned dire socio-economic outcomes including job losses as well as disruptions in supply of essential needs. Thus, immediate preventive, containment, and mitigation measures threaten to constitute “solutions” that are worse than the problem. These put the society in a dilemma between saving lives and saving the means of livelihood. This is where long term responses are called for.

As part of long-term response, economically advanced countries such as Germany and USA have already embarked on widespread, random antibodies testing for the SARS-CoV-2 virus. The US CDC has designed and validated a serologic test for broad-based surveillance and research that will provide information needed to guide the response to the pandemic and protect the public’s health^19^. Developing countries, on the other hand, face peculiar challenges in responding to COVID-19 both immediately and in the long term^20^. Otu et al. substantiate the question whether the African continent can play the long game of managing the combined public health and socio-economic impacts of COVID-19^20^. In a bid to gather information that will guide long term responses to COVID-

19 in sub-Saharan Africa, we have embarked on a seroepidemiologic survey of communities and populations in Cross River and Akwa Ibom States of Nigeria, beginning with this pilot study at a tertiary healthcare facility, the University of Calabar Teaching Hospital, UCTH. The results we present here indicate community spread within and beyond UCTH and call for immediate containment efforts by all and sundry, hence, the need to publish even with 66 participants tested. The seroepidemiological survey is ongoing.

## 2. RESEARCH METHODOLOGY

This study received ethical approval (UCTH/HREC/33/703) from the University of Calabar Teaching Hospital Health Research Ethics Committee. The main test kit used, namely, COVID-19 IgG/IgM Rapid Test Cassette (Whole Blood/Serum/Plasma) received emergency use authorization (EUA) from the FDA (EUA200056 Healgen LOA 05-29-2020) for “use as an aid in identifying individuals with an adaptive immune response to SARS-CoV-2, indicating recent or prior infection”. The test kit, henceforth denoted as FDA-EUA-Kit, was purchased from Confirm Biosciences (10123 Carroll Canyon Road, San Diego, CA 92131), the sole distributor of Healgen products in North America. Simultaneously with the FDA-EUA-Kit, another kit (manufactured by Zhejiang Orient Gene Biotech) used in 44 of the 66 total participants, was donated to UCTH by Lafarge Africa Plc. This kit will be denoted “Other Kit”, henceforth.

The initial 66 participants were randomly sampled from volunteering staff and patients of the UCTH from 17^th^ June to 25^th^ June 2020. They comprised of adults (>18 years of age), with 38 (58%) male and 28 (42%) female. Of the 66 participants, 62 (94%) had resided in Calabar Municipality or Calabar South Local Government Area of Cross River State for at least 12 months prior to the testing. Informed consent was obtained from all participants via signing of the Informed Consent Form (Supplementary Information, SI I, II) after explaining the objectives of the study to them. A structured questionnaire (SI III) administered to the participants by trained research assistants was used to capture personal data, demographic information, and past and present records of COVID-19 symptoms. SI IV shows inclusion and exclusion criteria for participation.

Details of blood collection and testing are given in SI V. About 5 μl of blood was collected in a capillary tube from a finger prick and this was preceded by disinfection using alcohol-based wipes. The specimen was dispensed into the well of the cassette and 2 drops of buffer solution was added. A timer was set to 10 minutes for FDA-EUA-Kits and 15 minutes for the other kits. After 10 or 15 minutes, test results were read. A total of three detection lines were possible, with the control (C) line appearing red when sample had flowed through the cassette. These were interpreted as shown in Figure 1, as per the manufacturer’s directives:

**Figure 1:**
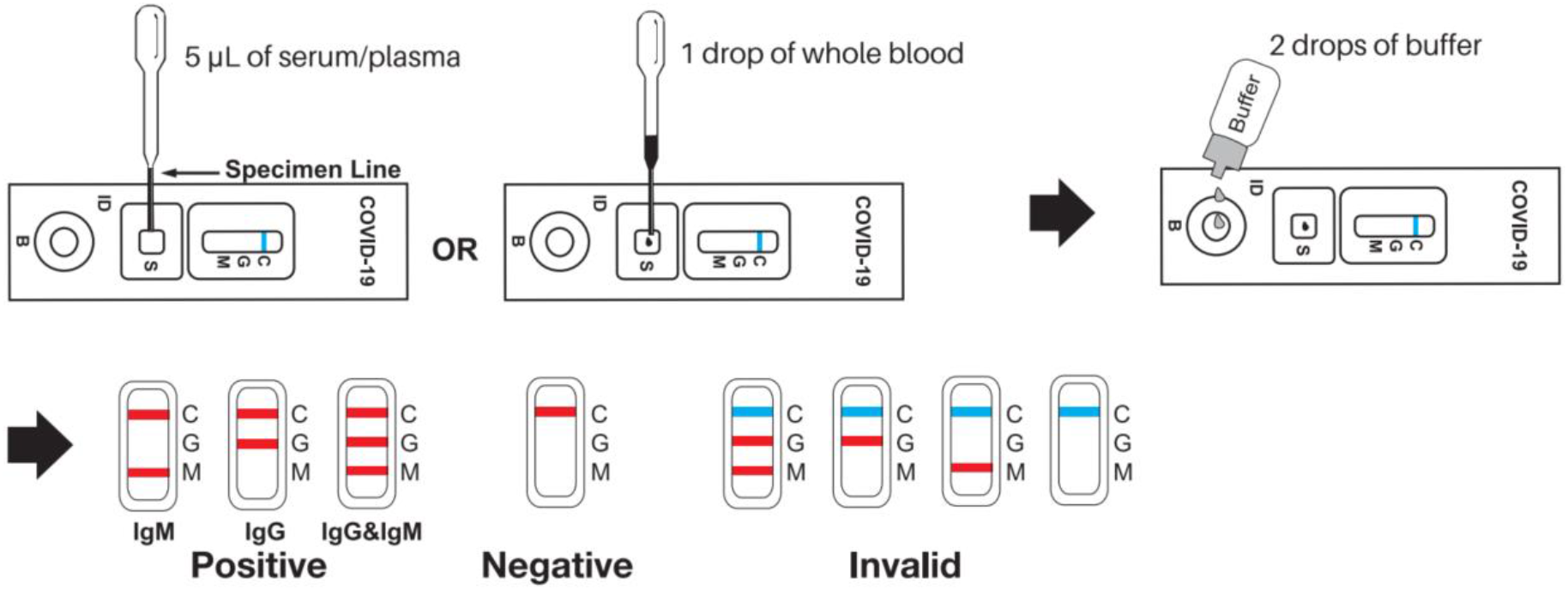
Testing steps and interpretation of COVID-19 antibody results (adapted from product insert, Healgen Scientific LLC, 3818 Fuqua St, Houston, TX 77047, USA).

1. Invalid Result: if the control line did not turn from blue to red, regardless of the other lines appearing or not, the test was deemed invalid.
2. Negative Result: if only the quality control line (C) appeared red and the detection lines G and M were not visible.
3. Positive Result, M only: if both the quality control line (C) and the detection line M appeared (red), the result was positive for the IgM antibody.
4. Positive Result, G only: if both the quality control line (C) and the detection line G appeared (red), then the novel coronavirus IgG antibody was positive for the IgG antibody.
5. Positive Result, G and M: if the quality control line (C) and both detection lines G and M appeared (red), then the result was positive for both the IgG and IgM antibodies.

Care was taken to place samples from each participant in both sets of test cassettes at the same time, with the results read from both after 10 or 15 minutes accordingly. Pictures of both cassettes placed on the consent forms of the participant were taken by the research assistants, as further evidence of test carried out, to eliminate any doubts over tabulated summary sheets as shown in Figure 2 for two participants.

**Figure 2:**
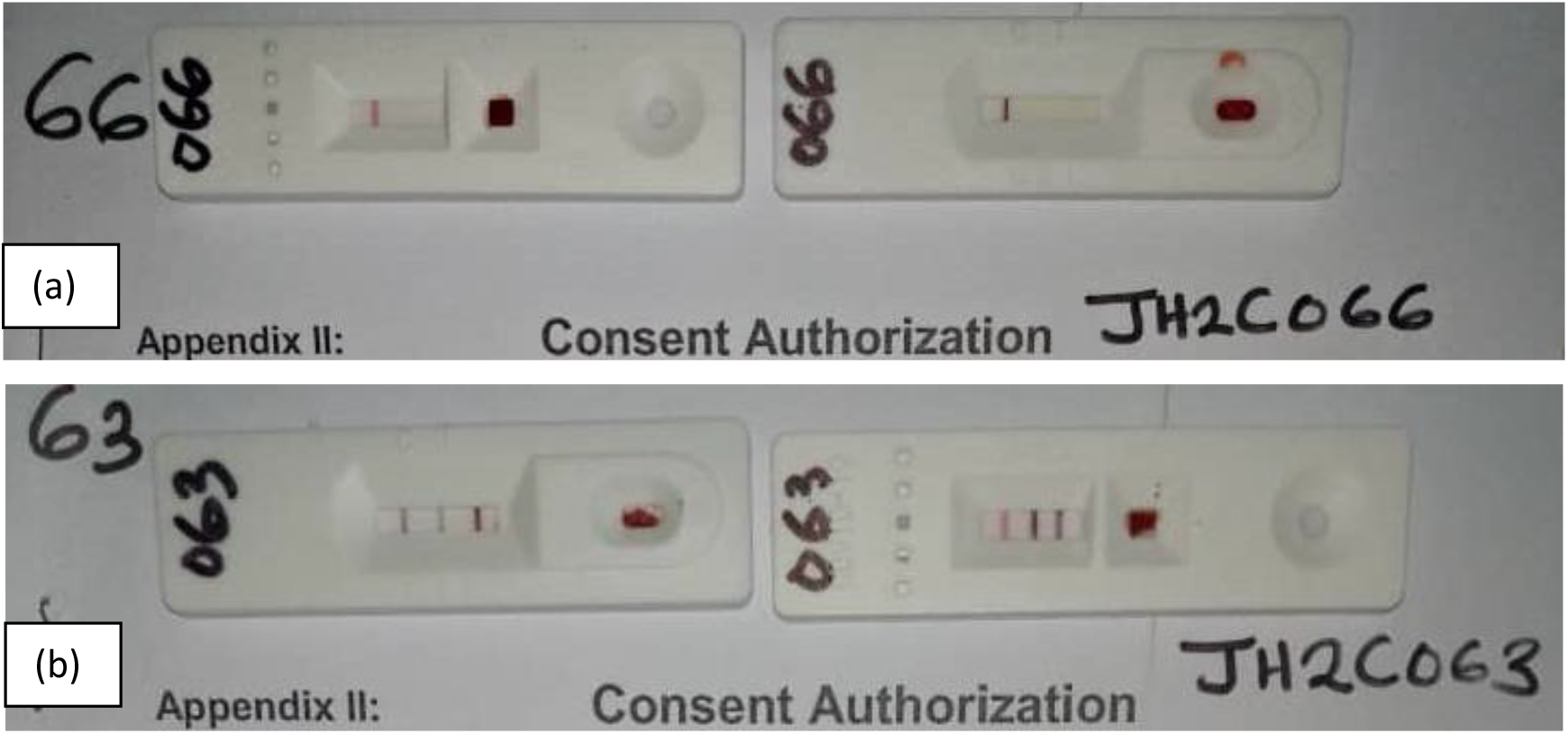
Pictures of test cassettes with results, on participants’ consent forms. (a) Both cassettes show negative results for participant 66. (b) Both cassettes show IgM and IgG positive for participant 63. In (b), the FDA-EUA-Kit is pictured on the right.

## 3. RESULTS

As shown in Figure 3, the FDA-EUA Kit test results for 5 of the 66 participants were both IgG and IgM positive (7.6%), 17 of the 66 were IgG positive (26%) while 49 (74%) were negative. There was no inconclusive result. For 44 of these 66 participants, simultaneous testing with the other kit yielded results that completely matched except one case where the FDA-EUA kit showed positive for both IgG and IgM while the other kit was positive only for IgM. Even for this one case, since the detection of IgM has the same implication as the detection of IgG and IgM, namely, acute or recent infection, the two test kits produced the same results. As shown in Figure 3, for the 44 participants tested with the “Other Kit” in addition to the FDA-EUA Kit, there were 4 IgG and IgM positive and 9 IgG positive. These results provide evidence of possible community transmission of COVID-19 in UCTH and Calabar, since 62 (94%) of participants had lived in Calabar for at least 12 months before the research. The work here serves as a pilot study to guide public health policies in response to COVID-19 pandemic both in the general population and in healthcare settings.

**Figure 3:**
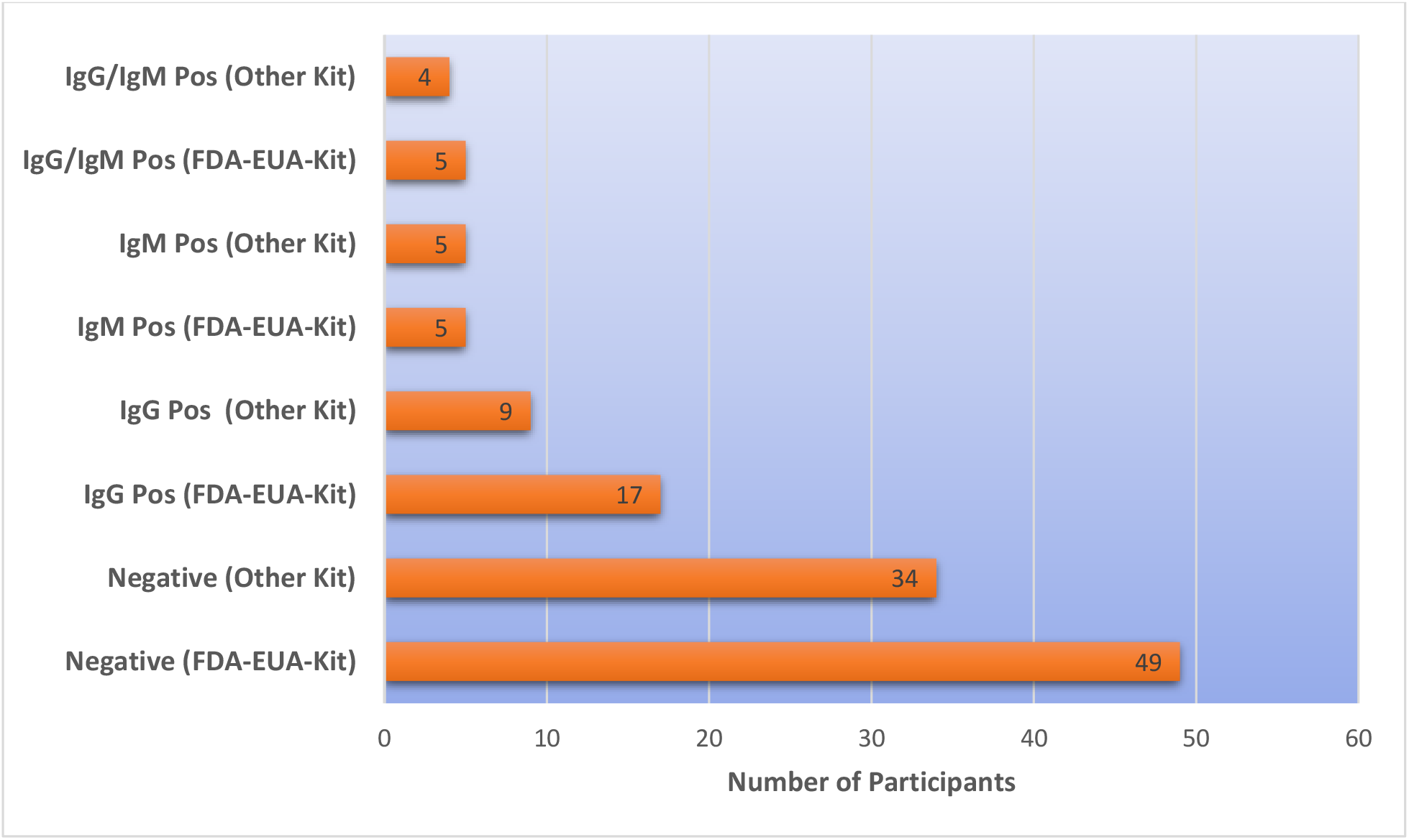
Prevalence of IgG and IgM antibodies to SARS-CoV-2. The FDA-EUA-Kits used for all 66 participants show 49 negative, 17 positive for IgG and 5 positive for both IgG and IgM. The “Other Kits” used for parallel testing of 44 out of the 66 participants show 34 negative, 9 IgG positive and 5 IgM positive (including 4 both IgM and IgG positive).

## 4. DISCUSSION

The fact that results from two test kits produced by different manufacturers completely overlapped for samples from all 44 participants tested with both kits increases confidence in the overall reliability of these results. The FDA-EUA-Kits showed 49/66 (74%) samples as negative for both IgM and IgG antibodies to SARS-CoV-2. The other kit showed 34/44 (77%) samples as negative for both IgM and IgG antibodies. Thus, participants tested were 76±1% negative for IgM and IgG antibodies, which implies that these individuals have either not been exposed to COVID-19 or the tests were done too early following infection, since it can take 1-3 weeks for antibodies to be developed following infection^15,21^ with the median being day 13^22^.

Although COVID-19 antibody tests are not designed to be the sole basis for diagnosing active infection^23^, validated test kits for the SARS-CoV-2 virus, such as the FDA-EUA Kit we used in our work, can indicate acute ongoing or recent infection when the result is IgM positive or both IgM and IgG positive. Thus, with 5 samples of the 66 participants in our study being both IgG and IgM positive (7.6%), there is evidence of ongoing infection in UCTH and Calabar. Even more worrisome epidemiologically is the fact that 17 of the 66 were IgG positive (26%) because that indicates recent COVID-19 infection, which means those participants have already spread the infection to others without knowing their status.COVID-19 IgG antibodies remain in the blood after an infection has passed. A serological testing for only IgG antibodies among clinical staff in a healthcare setting in Germany reported only 2.7% infection rate^24^. A seroepidemiological study that combined PCR testing with antibody testing in a German community found 15.5% infection rate, 5 times higher than the official PCR-only-based report of 3.1% for the same community^25^. Also using both PCR tests and IgG/IgM antibody tests for cohorts of healthcare workers, hospitalized patients and general workers in Wuhan, China, it was found that IgG prevalence was only 4.0% in healthcare providers and 4.6% in general workers^26^. While direct comparisons may not be completely warranted owing to our modest sample number of 66, the trend of lower infection rate among healthcare workers compared to others in these reports^24–26^ suggests that the infection rate in the community where UCTH is situated may even be higher than the 26% we have found. Thus, in spite of the relatively low case fatality rate in Nigeria based on Nigeria’s Center for Disease Control (NCDC) data^27^ which is about 2%, our result indicates the urgent need for enhanced diagnostic testing, contact tracing, isolation of infected individuals and more drastic measures to contain the pandemic in the community. This spurred us to bring these results of the pilot study to the public domain, while continuing with the larger scale seroepidemiological survey. Importantly, the combination of PCR testing with a serological survey presents some advantages that we highlight next.

In their vast review of current clinical practice and literature, Espejo et al.^22^ considered 55 recent reports on the use of serological testing and found that simultaneous IgG/IgM measurements significantly adds sensitivity to RT-PCR testing protocols early post-onset of symptoms and becomes the most accurate diagnostic test at later time points^22^. This is not surprising because RT-PCR sensitivity is subject to viral loads since it detects viral genetic material, and, in turn, viral loads may depend on the time of testing following infection and on sample collection. Although it is the gold standard, false negatives have been reported with RT-PCR, especially in the first week of infection^28^. A call for updated diagnostic clinical algorithm for COVID-19 in settings such as Nigeria where even RT-PCR is not always available for use during the most sensitive window for it is indeed timely^29^. Our results in this work add to the urgency of combined availability of RT-PCR and serological testing in such settings.

A good illustration of the utility of antibody testing in addition to PCR is the recent preliminary result for 3,000 New Yorkers randomly tested in which 13.9% were positive for coronavirus antibodies (about 21% in New York City itself)^30^, revealing a rate of infection much higher than RT-PCR diagnostic counts, thereby indicating a lower fatality rate for COVID-19. This lower fatality rate, also reported by many other countries based on antibody testing, more clearly explains the association between co-morbidities and COVID-19 fatality, allowing resources to be better allocated for mitigation of co-morbidities.

## 5. CONCLUSION

Current standard diagnostic tests for COVID-19 are relatively expensive and challenging to carry out on entire populations, even in advanced economies. The testing rate is crucial to interpreting case reports from various countries. Unless the testing rate is substantially increased, immediate responses to COVID-19 in Nigeria will remain inadequate and poorly informed. With poorly informed containment and mitigation strategies, long term responses are in even greater jeopardy. While the problem of knowing who is infected and who is not remains to be addressed through dedicated diagnostic testing, we have used serological testing as an easier means of addressing the question: to what extent has COVID-19 spread within a healthcare setting and within the community? Our preliminary results revealing 17/66 (26%) IgG positive and 5/66 (7.6%) IgM positive populations in UCTH, a tertiary health care facility, call for more diagnostic testing and more serological surveys of the general population in Cross River State and beyond as part of measures to contain COVID-19 pandemic.

## Data Availability

All data referred to in the paper is available upon request.

## ACKNOWLEDGEMENTS

The authors thank Prof Ekanem Braide for reading and editing the manuscript.

## AUTHOR CONTRIBUTIONS

Designed the study; MIA, VU, AO, OG, AI, SA, EE, AE, JG; performed the testing; MAI, ME, WE, UU, OI; analyzed data and plotted results; AE, MIA, AO, SA; wrote the manuscript; AE, AO, EE, BG, MIA; contributed equipment, testing kits, reagents; JG, MAI, AE; UU, OI.

